# Validation of a multi-gene qPCR-based pharmacogenomics panel across major ethnic groups in Singapore and Indonesia

**DOI:** 10.1101/2021.05.10.21256948

**Authors:** Anar Sanjaykumar Kothary, Caroline Mahendra, Mingchen Tan, Eunice Jia Min Tan, Jacelyn Phua Hong Yi, Gabriella, Tan Xin Hui Jocelyn, Jessline Stephanie Haruman, Zhihao Tan, Chun Kiat Lee, Alexander Lezhava, Benedict Yan, Astrid Irwanto

## Abstract

**Background:** With up to 70% of adverse drug reactions (ADRs) having high genetic associations, the clinical utility of pharmacogenomics (PGx) has been gaining traction. Nala PGx Core™ is a multi-gene qPCR-based panel that comprises 18 variants and 2 *CYP2D6* Copy Number markers across 4 pharmacogenes – *CYP2C9, CYP2C19, CYP2D6* and *SLCO1B1*.

**Objectives:** In this study, we validated the performance of Nala PGx Core™ against benchmark methods, on the Singaporean and Indonesian populations. Additionally, we examined the allele and diplotype frequencies across 5 major ethnic groups present in these populations namely, Indonesians, Chinese, Malays, Indians and Caucasians.

**Methods:** Human gDNA samples, extracted from the buccal swabs of 246 participants, were tested on Nala PGx Core™ and two chosen benchmarks, Agena VeriDose^®^ Core and *CYP2D6* Copy Number Variation (CNV) Panel, and TaqMan^®^ DME Genotyping Assays. Performance was evaluated based on assay robustness, precision and accuracy at the genotype- and diplotype-level.

**Results:** Nala PGx Core™ demonstrated high genotype- and diplotype-level call rates of >97% and >95% respectively in *CYP2D6*, and 100% for *CYP2C9, CYP2C19* and *SLCO1B1*. A precision rate of 100% was observed on both intra- and inter-precision studies. Variant-level concordance to the benchmark methods was ≥96.9% across all assays, which consequently resulted in a diplotype-level concordance of ≥94.7% across *CYP2C9, CYP2C19* and *CYP2D6*. Overall, the allele frequencies of *CYP2D6*10* and *CYP2D6*36* were higher in our cohort as compared to previous records. Notably, *CYP2D6* copy number variation (CNV) analysis demonstrated a *CYP2D6* *10/*36 frequency of 26.5% amongst the Indonesian cohort.

**Conclusion:** Nala PGx Core™ produced robust and accurate genotyping when compared to other established benchmarks. Furthermore, the panel successfully characterized alleles of clinical relevance in the Singaporean and Indonesian populations such as *CYP2D6*10* and *CYP2D6*36*, suggesting its potential for adoption in clinical workflows regionally.

## Introduction

Pharmacogenomics (PGx) is a branch of medicine concerned with determining the response of an individual to therapeutic drugs using genetic and/or genomic information^1^. With up to 70% of adverse drug reactions (ADRs) having high genetic associations^2^, and the sunk cost of trial-and-error prescriptions amounting to USD 30 billion^3^, the clinical utility^4^ of PGx appears to be gaining prominence.

Applications of PGx include the genotyping of Cytochrome P450 (P450) enzymes which are significantly involved in drug metabolism and chemical toxicity^5^. *CYP2C9, CYP2C19* and *CYP2D6* display common polymorphisms that produce altered enzyme activity, often resulting in deviant drug metabolism that leads to side effects from drugs metabolized by them^5^.

Alterations in *CYP2C9* can disrupt the metabolism of warfarin, while *CYP2C19* is associated with the metabolism of a number of drugs including clopidogrel, omeprazole and phenytoin^6,7^. The highly polymorphic *CYP2D6* has more than 100 variants and is responsible for the metabolism of approximately 25% of all clinically used drugs, including codeine, fluoxetine and amitriptyline^7^. For this reason, pharmaceuticals are particularly cautious in the development of drugs that are known to interact with these three P450s^5^. Additionally, pharmacogenes, such as *SLCO1B1*, are responsible for encoding transporters of simvastatin and other statins into tissues also affect their response upon intake in individuals. Variants of this gene are associated with increased risk of muscle pain (myalgia) and damage, including myopathy and in severe cases rhabdomyolysis^8^. Previous studies suggest the role of specific *CYP2C19* and *SLCO1B1* variants in the bioactivation of drugs such as clopidogrel and atorvastatin in the Singaporean population^9,10^. The *CYP2C19* variant, rs4986893, which is implicated in reduced clopidogrel metabolism, has been observed to be 50 times more prevalent in the Singaporean Chinese population than in Europeans^10^. The *SLCO1B1* variant, rs4149056, which is implicated in statin metabolism, has been observed to occur less frequently in the Singaporean population^9^. Taken together, these cases highlight the need to harness PGx testing to account for population differences in the personalization of prescriptions^11^.

Current methods on PGx testing for the identification of genetic polymorphisms revolve around variant genotyping that utilize technologies ranging from Next Generation Sequencing (NGS), Microarray, Mass Spectrometry to Quantitative PCR (qPCR)^1,6,12,13^. Main considerations in the adoption of a platform include its robustness, effectiveness and cost-saving, especially in localities with limited resources. qPCR-based assays satisfy these criteria and are a preferred platform for clinical laboratories^14–19^. However, some of the available commercial kits are limited in their ability to test the 4 genes of interest mentioned above in a single panel^20–25^. Nala PGx Core™ is a qPCR-based multi-gene panel consisting of 20 pharmacogenomic variants that are associated with altered *CYP2C9, CYP2C19, CYP2D6* and *SLCO1B1* activity in Asian geographies. Our study aims to evaluate the performance of Nala PGx Core™ in effectively identifying genetic variations that may contribute to PGx-related clinical outcomes in Singapore and Indonesia, with a secondary objective of highlighting the allele and diplotype frequencies in the geographies examined.

## Methods and Materials

### Ethics Approval

Institutional Review Board (IRB) approval was granted by the Parkway Independent Ethics Committee (Singapore) under IRB Reference Number PIEC/2019/021 and Siloam Hospitals Ethics Review Committee (Jakarta, Indonesia) under IRB Reference Number 001/EA/KEPKK/RSMRCCC/V/2019.

### Study Recruitment

Patients were recruited on behalf of Nalagenetics Pte. Ltd. with written informed consent forms from recruitment sites in Singapore and Indonesia. A total of 251 samples were evaluated from 5 major ethnic groups to ensure the representation of the major ethnic groups residing in both countries – Chinese, Malays, Indians, Caucasians and Indonesians. Participants identifying as one or more of the following ethnicities were categorized as Indonesians: Ambon, Batak, Betawi, Jawa, Lampung, Manado, Minangkabau, Nusa Tenggara Timur, Palembang, Sulawesi, Sunda, Timor Leste, Tolaki and Toraja.

Buccal samples were collected using OraCollect (Cat No. DNA OCR-100 from DNA Genotek) and genomic DNA (gDNA) extracted using the Monarch Genomic DNA Purification Kit (Cat No. T3010 from NEB). The extraction procedure followed manufacturer’s instructions with additional dry-spin step at maximum speed for 1 minute after the 2^nd^ buffer washing step. The quality and concentration of gDNA extracts were quantified by NanoDrop 2000 Spectrophotometer (Singapore) and BioDrop-µLITE (Indonesia). Samples that failed to meet the DNA quality control criteria (n=5) of A260/280 >1.7 and DNA yield >500ng were excluded from the study. The remaining extracted gDNA samples (n=246) were stored at -20□ for downstream application.

### Nala PGx Core™

The Nala PGx Core™ kit from Nalagenetics Pte. Ltd. consists of 20 qPCR-based variant assays across four genes – *CYP2C9, CYP2C19, CYP2D6* and *SLCO1B1*. Whilst assays for *CYP2C9, CYP2C19* and *CYP2D6* have been designed to enable the detection of specific star alleles, the *SLCO1B1* assay has been designed to detect the variant rs4149056, which is present in three reduced function haplotypes namely, *SLCO1B1*5, SLCO1B1*15* and *SLCO1B1*17*. The *SLCO1B1* assay is thus, unable to differentiate between each of the three aforementioned haplotypes. The variants covered by the kit are outlined in Table 1.

**Table 1.** Genes and variants evaluated

| Gene | Star Allele | Variant | Nucleotide Changes | Effect On Protein | Allele Clinical Function Status | Genotyping Methods Utilized |  |  |
| --- | --- | --- | --- | --- | --- | --- | --- | --- |
|  |  |  |  |  |  | Nala PGx Core™ | Agena VeriDose® Core and CYP2D6 CNV Panel | TaqMan® DME Genotyping Assays |
| CYP2C9 | *2 | rs1799853 | 3608C>T | R144C | Decreased | Bi-allelic Assay | VeriDose Core |  |
|  | *3 | rs1057910 | 42614A>C | I359L | None | Bi-allelic Assay | VeriDose Core |  |
| CYP2C19 | *2 | rs4244285 | 19154G>A | Splicing Defect | None | Bi-allelic Assay | VeriDose Core |  |
|  | *3 | rs4986893 | 17948G>A | W212X | None | Bi-allelic Assay | VeriDose Core |  |
|  | *17 | rs12248560 | -806C>T | 5' Region | Increased | Bi-allelic Assay | VeriDose Core |  |
| CYP2D6 | *2 | rs1135840 | 4181G>C | S486T | Normal | Bi-allelic Assay | VeriDose Core |  |
|  | *2 | rs16947 | 2851C>T | R296C | Normal | Bi-allelic Assay | VeriDose Core |  |
|  | *3 | rs35742686 | 2550delA | Frameshift | None | Bi-allelic Assay | VeriDose Core |  |
|  | *4 | rs3892097, rs1065852 | 1847G>A, 100C>T | Splicing Defect, P34S | None | Bi-allelic Assay | VeriDose Core |  |
|  | *5 | N/A | N/A | Gene Deletion | None | CNV Assay (Intron 2) | CYP2D6 CNV |  |
|  | *6 | rs5030655 | 1708delT | Frameshift | None | Bi-allelic Assay | VeriDose Core |  |
|  | *8 | rs5030865 | 1759G>T | G169X | None | Tri-allelic Assay | VeriDose Core |  |
|  | *9 | rs5030656 | 2616delAAG | K281del | Decreased | Bi-allelic Assay | VeriDose Core |  |
|  | *10 | rs3892097, rs1065852 | 1847G>A, 100C>T | Splicing Defect, P34S | Decreased | Bi-allelic Assay | VeriDose Core |  |
|  | *14 | rs5030865 | 1759G>A | G169R | Decreased | Tri-allelic Assay | VeriDose Core |  |
|  | *21 | rs72549352 | 2580_2581ins C | Frameshift | None | Bi-allelic Assay | VeriDose Core |  |
|  | *29 | rs59421388 | 3184G>A | V338M | Decreased | Bi-allelic Assay | VeriDose Core |  |
|  | *31 | rs267608319 | 4043G>A | R440H | None | Bi-allelic Assay | NA | AH21B9N |
|  | *35 | rs769258 | 31G>A | V11M | Normal | Bi-allelic Assay | NA | C__27102444_F0 |
|  | *36 | N/A | Recombination at Exon 9 | CYP2D6-2D7 Hybrid | None | CNV Assay (Exon 9) | CYP2D6 CNV |  |
|  | *41 | rs28371725 | 2989G>A | Splicing Defect | Decreased | Bi-allelic Assay | VeriDose Core |  |
| SLCO1B1 | NA<sup>†</sup> | rs4149056 | g.52422T>C | V174A | Decreased | Bi-allelic Assay | VeriDose Core |  |

<sup>†</sup>Nala PGx Core™ detects the variant, rs4149056, which is associated with decreased enzymatic activity and is present in three known *SLCO1B1* haplotypes namely, *SLCO1B1*\*5, *SLCO1B1*\*15 and *SLCO1B1*\*17.

Assays were set up on a 96-well plate. Human gDNA was added at a concentration of 2 ng/µL as template for the qPCR reaction, which was then performed on the Bio-Rad CFX96 IVD Touch™ Real-Time PCR Detection System per the product insert. Run analysis was performed using the application CFX Manager 3.1 or CFX Maestro, and exported as raw .csv files. Exported files were uploaded into the companion software, Nala Clinical Decision Support™ (Nala CDS™) for further analysis of variant genotyping, diplotype determination and phenotype translation. Genotyping using Nala PGx Core™ was performed at the Molecular Diagnosis Centre, National University Health System, Singapore (NUHS MDC) and PT Nalagenetik Riset Indonesia.

### Agena VeriDose^®^ Core and *CYP2D6* Copy Number Variation (CNV) Panel

The VeriDose^®^ Core and *CYP2D6* Copy Number Variation (CNV) Panel from Agena Bioscience^®^ consists of 68 variant assays in 20 genes and 5 *CYP2D6* CNV assays, accompanied by a reporting software that automatically analyzes each variation^26^. Genotyping using Agena VeriDose Core and *CYP2D6* CNV Panel was performed at the Genome Institute of Singapore. Variants evaluated using this platform are listed in Table 1. The Agena VeriDose^®^ Panel has been utilized by the United States Centers for Disease Control and Prevention (CDC) as part of their Genetic Testing Reference Material (GeT-RM) Coordination Program^27^.

### TaqMan^®^ Drug Metabolism Enzyme (DME) Genotyping Assay

TaqMan^®^ DME Genotyping Assays were utilized in the evaluation of *CYP2D6* rs769258 (TaqMan Assay ID AH21B9N) and *CYP2D6* rs267608319 (TaqMan Assay ID C_27102444_F0). Assays were set up on a 384-well plate^28^ with a sample input of human gDNA at 2 ng/µL. The subsequent PCR reaction was performed on the Applied Biosystems ViiA™ 7 Real-Time PCR System as per the recommended cycling conditions, at the Genome Institute of Singapore. Post-PCR plate read was performed using the companion software, TaqMan^®^ Genotyper™ Software for single nucleotide polymorphisms (SNP) genotyping. Similar to the Agena VeriDose^®^ Panel, TaqMan^®^ DME Genotyping Assays were employed in the characterization of DNA samples as part of the CDC GeT-RM program^27^.

### Robustness

Genotype- and diplotype-level call rates were defined as the percentage of samples that returned a genotype at the variant-level or were assigned a distinct diplotype for the gene of interest, respectively. Failed tests were defined as samples that did not return a genotype and/or diplotype call for the genes evaluated.

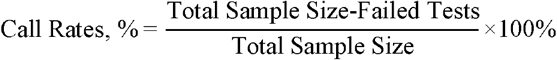

### Precision

Three samples at 3 DNA concentrations were tested across 3 reagent lots on 2 machines. Each test condition was repeated within the same plate for a triplicate. For variant assays that identified SNPs and indels, intra-precision was performed within the same plate, run as triplicates across 47 tests. Inter-precision was assessed from 120 tests performed across plate runs covering the 4 variables – samples, DNA concentration, reagent lots and machines. Concordance rates across precision studies were calculated as the percentage of tests that returned a genotype call concordant to the expected truth for each variant assay. Discordant genotype was defined as instances when the test returned a genotype call that was different from the expected truth.

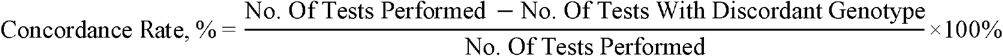

For *CYP2D6* CNV assays, copy number estimates for Intron 2 and Exon 9 of the three samples were derived based on their cycle threshold (Ct) results across plate runs.

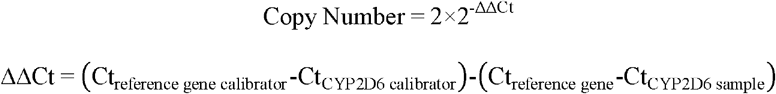

Testing of the three samples was repeated for a number of plate runs, n (Table 5), and calculated for the average copy number of each sample and their coefficient of variation (CV). The CV for each plate run was calculated by finding the standard deviation (*σ*_*plate*_) between triplicates within the same plate run, and divided by the triplicate mean (*μ*_*plate*_). The average of the individual CVs was reported as the intra-precision CV. For inter-precision CV, standard deviation population (*σ*_*plate means*_) was divided by the mean population, i.e. average of means.

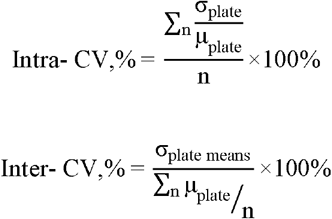

### Accuracy

#### Variant-level Concordance

The accuracy of Nala PGx Core™ in genotyping at a variant-level was evaluated by comparing calls produced by Nala PGx Core™ assay against benchmark methods as listed in Table 1. Samples that successfully produced genotype calls for all variants tested on Nala PGx Core™ and its benchmarks were considered for the evaluation (n = 225 for all variants except *CYP2D6* CNV; n = 224 for *CYP2D6* CNV). Samples that failed to produce a genotype call on one or more of the platforms were excluded from the concordance calculation (n = 21 for all variants except *CYP2D6* CNV; n = 22 for *CYP2D6* CNV, Supplementary Table 7). Discordant calls were defined as instances in which Nala PGx Core™ provided a genotype call that was different from that of a call made by the corresponding benchmark. Percentage concordance to the benchmark was calculated per variant as follows –

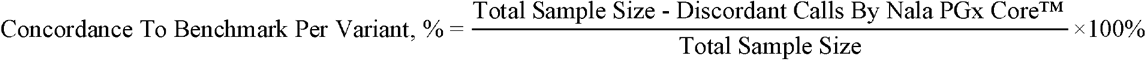

#### Diplotype-level Concordance

The accuracy of Nala PGx Core™ in assigning a diplotype call for *CYP2C9, CYP2C19*, and *CYP2D6*, was evaluated by comparing calls against the Agena VeriDose^®^ Core and *CYP2D6* CNV Panel. Samples that met the following criteria were included in the sample size of each gene:

1. Successful genotype-level calls on the relevant platforms for all variants covered by the gene of interest
2. Successful assignment of a diplotype for the gene of interest on both Nala PGx Core™, and Agena VeriDose^®^ Core and *CYP2D6* CNV Panel

Discordant calls were defined as instances in which Nala PGx Core™ assigned a diplotype that differed from the call made by the Agena VeriDose^®^ Core and *CYP2D6* CNV Panel.

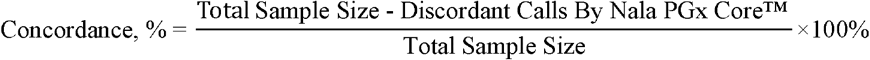

### Frequencies By Ethnicity

Ethnicities were obtained based on participant self-identification across both the population cohorts as part of the recruitment questionnaire. Participants that did not report an ethnic group in the recruitment form were excluded from the frequency analysis. The following were further excluded from the frequency analysis:

1. Samples in which participants did not report an ethnic group on the recruitment form (n=6)
2. Samples with one or more variant level failures across the 4 genes evaluated in Table 1 (n=18)
3. Samples with one or more diplotype-level failures (“No Call”) across *CYP2C9, CYP2C19*, and *CYP2D6* on Nala PGx Core™ and the Agena VeriDose^®^ Core and *CYP2D6* CNV Panel (n=15)
4. Samples with discordant diplotype calls for the gene of interest (n=variable, Supplementary Table 7)

The remaining samples were included in the allele-level frequency analysis of *CYP2C9* (n=206), *CYP2C19* (n=201), *CYP2D6* (n=195) and *SLCO1B1* (n=203), as well as in the diplotype-level frequency analysis of *CYP2C9* (n=206), *CYP2C19* (n=201) and *CYP2D6* (n=195). Allele and diplotype frequency values were derived using the following formulae^29,30^, for both the overall study cohort as well as for each ethnic group.

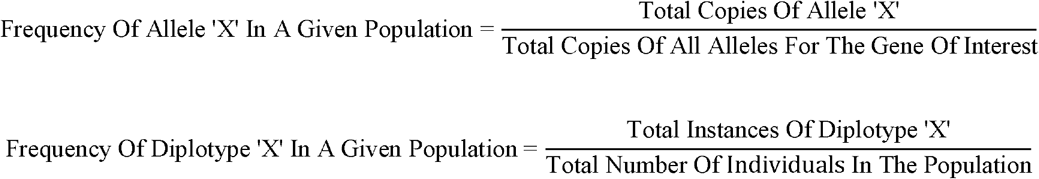

## Results

### Robustness

We first sought to evaluate the observed genotype- and diplotype-level call rates of the platforms evaluated in this study. 246 samples underwent variant genotyping and diplotype determination, across the four genes evaluated on the genotyping platforms (Tables 2, 3). The genotype-level call rates for Nala PGx Core™ were at 100% for *CYP2C9, CYP2C19* and *SLCO1B1*, and the diplotype-level call rates were at 100% for *CYP2C9* and *CYP2C19*. The benchmark platform, Agena VeriDose^®^ Core Panel, demonstrated call rates of >95.9% at the genotype-level and >90.7% at the diplotype-level.

**Table 2.** Observed genotype-level call rates per variant per gene per platform

| Gene | Variant | Variant Call Rate, % (n=246) |  |  |
| --- | --- | --- | --- | --- |
|  |  | Nala PGx Core™ | Agena VeriDose® Core and CYP2D6 CNV Panel | TaqMan® DME Genotyping Assays |
| CYP2C9 | rs1799853 | 100.0 | 99.2 | NA |
|  | rs1057910 | 100.0 | 99.6 | NA |
| CYP2C19 | rs4244285 | 100.0 | 99.6 | NA |
|  | rs4986893 | 100.0 | 99.6 | NA |
|  | rs12248560 | 100.0 | 98.8 | NA |
| CYP2D6 | rs1065852 | 98.4 | 95.9 | NA |
|  | rs5030655 | 100.0 | 99.2 | NA |
|  | rs3892097 | 98.8 | 99.2 | NA |
|  | rs35742686 | 100.0 | 98.8 | NA |
|  | rs16947 | 100.0 | 99.6 | NA |
|  | rs28371725 | 100.0 | 99.2 | NA |
|  | rs1135840 | 100.0 | 99.6 | NA |
|  | rs769258 | 98.8 | NA | 100.0 |
|  | rs5030865 | 97.2 | 99.2 | NA |
|  | rs5030656 | 100.0 | 99.6 | NA |
|  | rs59421388 | 100.0 | 99.2 | NA |
|  | rs267608319 | 99.6 | NA | 100.0 |
|  | CNV Assay (Intron 2) | 99.6 | 99.2 | NA |
|  | CNV Assay (Exon 9) | 99.6 | 99.2 | NA |
| SLCO1B1 | rs4149056 | 100.0 | 99.2 | NA |

**Table 3.**
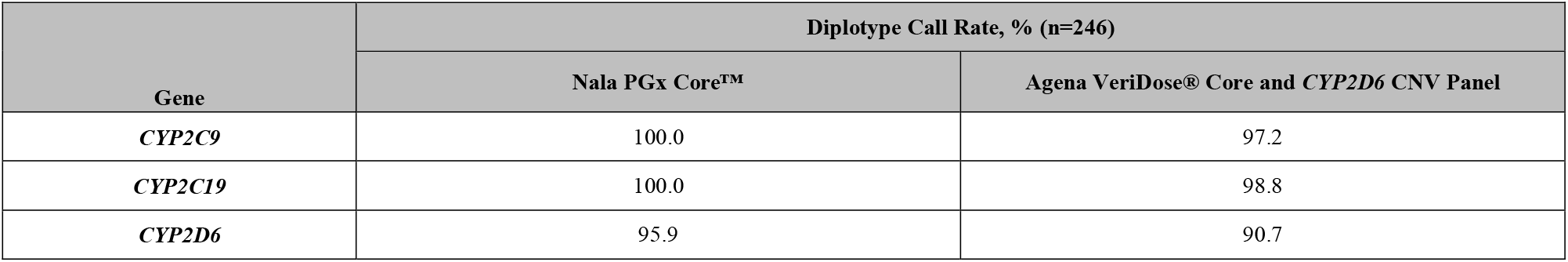
Observed diplotype-level call rates per gene per platform

| Gene | Diplotype Call Rate, % (n=246) |  |
| --- | --- | --- |
|  | Nala PGx Core™ | Agena VeriDose® Core and CYP2D6 CNV Panel |
| CYP2C9 | 100.0 | 97.2 |
| CYP2C19 | 100.0 | 98.8 |
| CYP2D6 | 95.9 | 90.7 |

Most variants in *CYP2D6*, except for seven, achieved 100% call rates on Nala PGx Core™, while the corresponding call rates of the benchmark platforms were observed to be between 95.9 – 99.2% on Agena VeriDose^®^ Core and *CYP2D6* CNV Panel, and 100% on TaqMan^®^ DME Genotyping Assays. Out of the seven aforementioned variants, Nala PGx Core™ demonstrated higher call rates than the benchmark for the genotyping of rs1065852, Intron 2 and Exon 9 variants. For rs3892097, rs769258, rs5030865, and rs267608319, the accompanying benchmarks demonstrated higher call rates. At the diplotype-level, Nala PGx Core™ demonstrated a CYP2D6 call rate of 95.9% as compared to the benchmark, which was observed to be at 90.7% (Table 3).

### Precision

A precision study was conducted to assess the consistency of Nala PGx Core™ for samples tested under the same conditions (intra-precision) and under different conditions (inter-precision). Both study resulted in 100% concordance for all assays across replicates, demonstrating consistent genotyping results across a range of DNA concentration, reagent lots and machine variations (Table 4). Precision of *CYP2D6* CNV assay was reported as the average copy number obtained for Intron 2 and Exon 9 of three samples, and their CV calculated across the test conditions (Table 5). The intra-CV ranged from 3-6% while inter-CV between 5-13%, demonstrating high precision of the assays across variables, where acceptable ranges were intra-CV below 10% and inter-CV below 15%^31^.

**Table 4.** Intra-precision and inter-precision concordance rates

| Gene | Variant | Concordance rate, % |  |
| --- | --- | --- | --- |
|  |  | Intra-precision<br>(47 tests in triplicate) | Inter-precision<br>(120 tests across 4 variables) |
| CYP2C9 | rs1799853 | 100% | 100% |
|  | rs1057910 | 100% | 100% |
| CYP2C19 | rs4244285 | 100% | 100% |
|  | rs4986893 | 100% | 100% |
|  | rs12248560 | 100% | 100% |
| CYP2D6 | rs1065852 | 100% | 100% |
|  | rs5030655 | 100% | 100% |
|  | rs3892097 | 100% | 100% |
|  | rs35742686 | 100% | 100% |
|  | rs16947 | 100% | 100% |
|  | rs28371725 | 100% | 100% |
|  | rs1135840 | 100% | 100% |
|  | rs769258 | 100% | 100% |
|  | rs5030865 | 100% | 100% |
|  | rs5030656 | 100% | 100% |
|  | rs59421388 | 100% | 100% |
|  | rs267608319 | 100% | 100% |
| **SLCO1B1** | rs4149056 | 100% | 100% |

**Table 5.** Intra-precision and inter-precision for *CYP2D6* Copy Number

| Sample | Variant | Average Copy Number | Sample Size, n | Coefficient of Variation |  |
| --- | --- | --- | --- | --- | --- |
|  |  |  |  | Intra-precision | Inter-precision |
| 221374 | Intron 2 | 2 | 11 | 4% | 6% |
|  | Exon 9 | 1 |  | 5% | 5% |
| 222660 | Intron 2 | 2 | 9 | 3% | 5% |
|  | Exon 9 | 2 |  | 4% | 6% |
| 222769 | Intron 2 | 3 | 22 | 4% | 10% |
|  | Exon 9 | 2 |  | 6% | 13% |

### Accuracy

#### Variant-level Concordance

To assess the accuracy of the panel, 18 SNPs and 2 *CYP2D6* Copy Number assays were genotyped on the panel, Nala PGx Core™, against benchmark methods as listed in Table 1. The 225 sample cohort consisted of DNA samples isolated from buccal swabs that had successfully produced genotype calls for all variants tested on Nala PGx Core™ and its benchmarks. The sample size per genotype displayed in Table 6 refers to the genotype calls made by the benchmark.

**Table 6.**
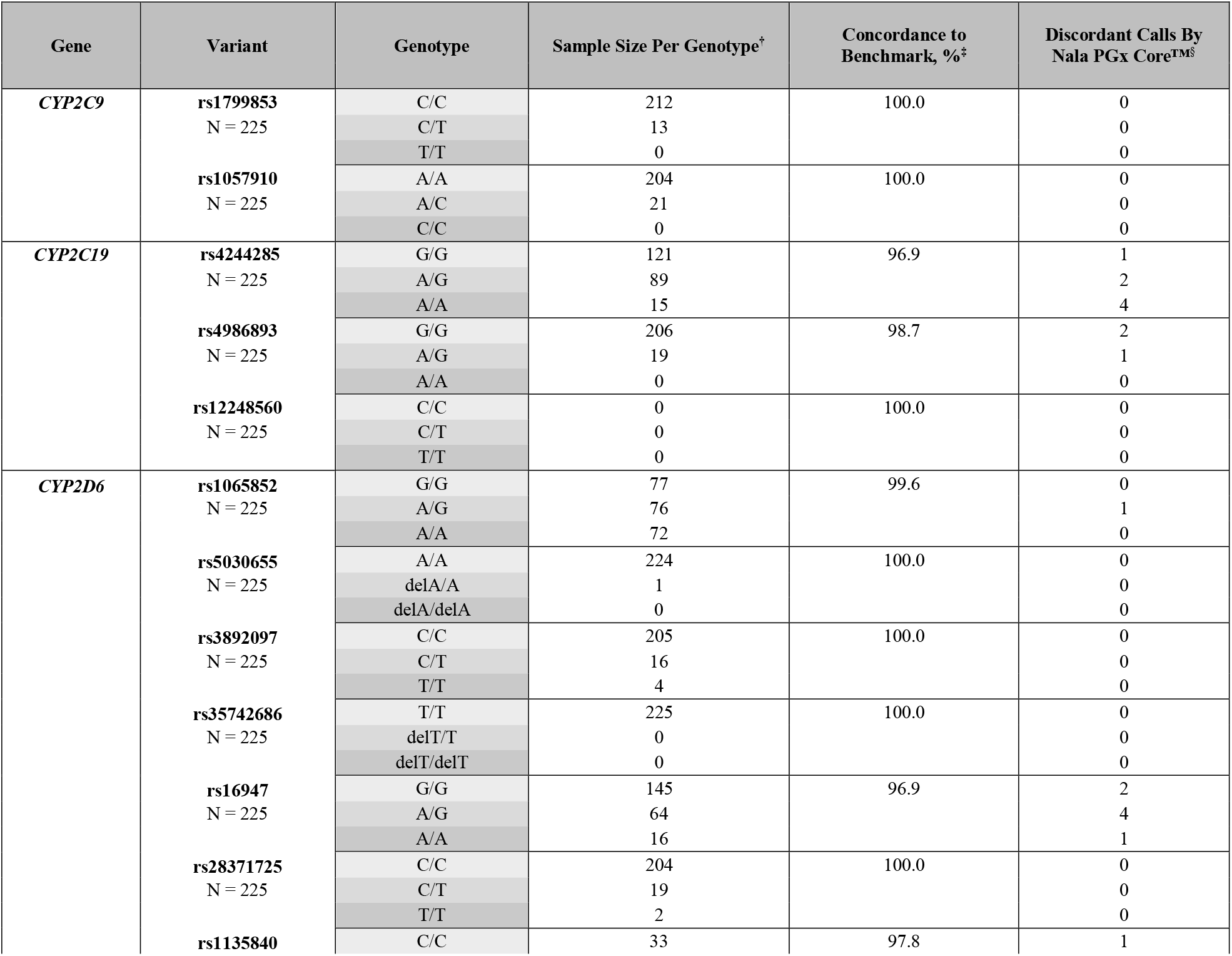
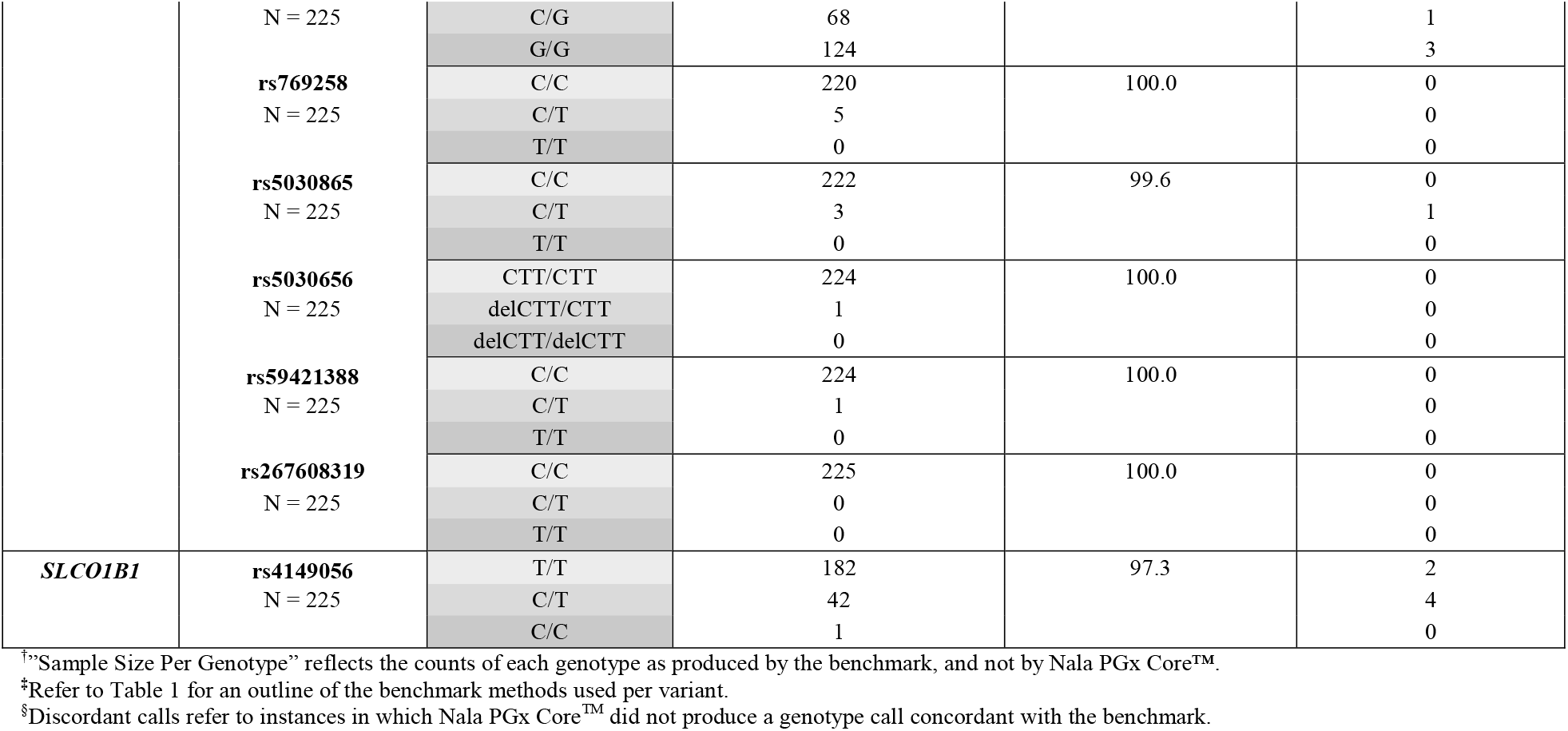
Genotype concordance for genes *CYP2C9, CYP2C19, CYP2D6* and *SLCO1B1*

| Gene | Variant | Genotype | Sample Size Per Genotype<sup>†</sup> | Concordance to Benchmark, %<sup>‡</sup> | Discordant Calls By Nala PGx Core™<sup>§</sup> |
| --- | --- | --- | --- | --- | --- |
| *CYP2C9* | rs1799853<br>N = 225 | C/C | 212 | 100.0 | 0 |
|  |  | C/T | 13 |  | 0 |
|  |  | T/T | 0 |  | 0 |
|  | rs1057910<br>N = 225 | A/A | 204 | 100.0 | 0 |
|  |  | A/C | 21 |  | 0 |
|  |  | C/C | 0 |  | 0 |
| *CYP2C19* | rs4244285<br>N = 225 | G/G | 121 | 96.9 | 1 |
|  |  | A/G | 89 |  | 2 |
|  |  | A/A | 15 |  | 4 |
|  | rs4986893<br>N = 225 | G/G | 206 | 98.7 | 2 |
|  |  | A/G | 19 |  | 1 |
|  |  | A/A | 0 |  | 0 |
|  | rs12248560<br>N = 225 | C/C | 0 | 100.0 | 0 |
|  |  | C/T | 0 |  | 0 |
|  |  | T/T | 0 |  | 0 |
| *CYP2D6* | rs1065852<br>N = 225 | G/G | 77 | 99.6 | 0 |
|  |  | A/G | 76 |  | 1 |
|  |  | A/A | 72 |  | 0 |
|  | rs5030655<br>N = 225 | A/A | 224 | 100.0 | 0 |
|  |  | delA/A | 1 |  | 0 |
|  |  | delA/delA | 0 |  | 0 |
|  | rs3892097<br>N = 225 | C/C | 205 | 100.0 | 0 |
|  |  | C/T | 16 |  | 0 |
|  |  | T/T | 4 |  | 0 |
|  | rs35742686<br>N = 225 | T/T | 225 | 100.0 | 0 |
|  |  | delT/T | 0 |  | 0 |
|  |  | delT/delT | 0 |  | 0 |
|  | rs16947<br>N = 225 | G/G | 145 | 96.9 | 2 |
|  |  | A/G | 64 |  | 4 |
|  |  | A/A | 16 |  | 1 |
|  | rs28371725<br>N = 225 | C/C | 204 | 100.0 | 0 |
|  |  | C/T | 19 |  | 0 |
|  |  | T/T | 2 |  | 0 |
|  | rs1135840 | C/C | 33 | 97.8 | 1 |
|  | N = 225 | C/G | 68 |  | 1 |
|  |  | G/G | 124 |  | 3 |
|  | **rs769258**<br>N = 225 | C/C | 220 | 100.0 | 0 |
|  |  | C/T | 5 |  | 0 |
|  |  | T/T | 0 |  | 0 |
|  | **rs5030865**<br>N = 225 | C/C | 222 | 99.6 | 0 |
|  |  | C/T | 3 |  | 1 |
|  |  | T/T | 0 |  | 0 |
|  | **rs5030656**<br>N = 225 | CTT/CTT | 224 | 100.0 | 0 |
|  |  | delCTT/CTT | 1 |  | 0 |
|  |  | delCTT/delCTT | 0 |  | 0 |
|  | **rs59421388**<br>N = 225 | C/C | 224 | 100.0 | 0 |
|  |  | C/T | 1 |  | 0 |
|  |  | T/T | 0 |  | 0 |
|  | **rs267608319**<br>N = 225 | C/C | 225 | 100.0 | 0 |
|  |  | C/T | 0 |  | 0 |
|  |  | T/T | 0 |  | 0 |
| **SLCO1B1** | **rs4149056**<br>N = 225 | T/T | 182 | 97.3 | 2 |
|  |  | C/T | 42 |  | 4 |
|  |  | C/C | 1 |  | 0 |

<sup>†</sup>“Sample Size Per Genotype” reflects the counts of each genotype as produced by the benchmark, and not by Nala PGx Core™.
<sup>‡</sup>Refer to Table 1 for an outline of the benchmark methods used per variant.
<sup>§</sup>Discordant calls refer to instances in which Nala PGx Core™ did not produce a genotype call concordant with the benchmark.

11 variants (*CYP2C9* rs1799853, rs1057910; *CYP2C19* rs12248560; *CYP2D6* rs5030655, rs3892097, rs35742686, rs28371725, rs769258, rs5030656, rs59421388, rs267608319) were genotyped against Agena VeriDose^®^ Core with a resulting concordance rate of 100% (N = 225 samples). Discordance was observed for *CYP2C19* rs4244285 (n=7), *CYP2C19* rs4986893 (n=3), *CYP2D6* rs1065852 (n=1), *CYP2D6* rs16947 (n=7), *CYP2D6* rs1135840 (n=5), *CYP2D6* rs5030865 (n=1) and *SLCO1B1* rs4149056 (n=6), with a mismatch rate of 0.44% to 3.1% for the affected assays. Overall, Nala PGx Core™ demonstrated >96% concordance to the benchmark, Agena VeriDose^®^ Core, for the 16 variants across 225 samples (Table 6).

Variants not present on Agena VeriDose^®^ Core, *CYP2D6* rs769258 and *CYP2D6* rs267608319, were genotyped using TaqMan^®^ DME Genotyping Assays. Nala PGx Core™ demonstrated 100% concordance (N = 225) to the benchmark for both SNPs.

For the *CYP2D6* Intron 2 and Exon 9 Copy Number assays, concordance was observed to be at 99.6% and 98.7% respectively, against the Agena *CYP2D6* CNV Panel. Discordant calls were observed in samples with an Intron 2 copy number greater than 3 (n=1), and for samples with an Exon 9 copy number of one (n=1) and two (n=2). The sample size per copy number displayed in Table 7 refers to the copy number calls made by the benchmark.

**Table 7.** Genotype concordance for *CYP2D6* Intron 2 And Exon 9 Copy Number Variations

| Gene | Variant | Copy Number | Sample Size Per Copy Number<sup>†</sup> | Concordance to Benchmark, %<sup>‡</sup> | Discordant Calls By Nala PGx Core™<sup>§</sup> |
| --- | --- | --- | --- | --- | --- |
| *CYP2D6* | **Intron 2**<br>N = 224 | 0 | 0 | 99.6 | 0 |
|  |  | 1 | 12 |  | 0 |
|  |  | 2 | 130 |  | 0 |
|  |  | >=3 | 82 |  | 1 |
|  | **Exon 9**<br>N = 224 | 0 | 0 | 98.7 | 0 |
|  |  | 1 | 24 |  | 1 |
|  |  | 2 | 193 |  | 2 |
|  |  | >=3 | 7 |  | 0 |

<sup>†</sup>“Sample Size Per Copy Number” reflects the counts of each copy number call as produced by the benchmark, and not by Nala PGx Core™.
<sup>‡</sup>Refer to Table 1 for an outline of the benchmark methods used per variant.
<sup>§</sup>Discordant calls refer to instances in which Nala PGx Core™ did not produce a copy number call concordant with the benchmark.

A summary of the concordance percentage as well as the distribution of genotypes of each variant are displayed in Table 6. Similarly, the distribution of *CYP2D6* Intron 2 and Exon 9 copy numbers are displayed in Table 7.

#### Diplotype-level Concordance

Following successful genotyping at the variant level, we investigated the accuracy of Nala PGx Core™ in assigning a diplotype call for *CYP2C9, CYP2C19* and *CYP2D6*, with reference to the Agena VeriDose^®^ Core and *CYP2D6* CNV Panel. Table 8 displays the percentage concordance after the further exclusion of samples that demonstrated diplotype mismatches arising from technological differences, where technological differences refer to the varying allele coverage of each platform (Supplementary Table 7). These differences were derived from the variant lists of both Nala PGx Core™ (Table 1) and its benchmark, the Agena VeriDose^®^ Core and *CYP2D6* CNV Panel^26^.

**Table 8.** Diplotype concordance for *CYP2C9, CYP2C19* and *CYP2D6* between Nala PGx Core™, and Agena VeriDose^®^ Core and *CYP2D6* CNV Panel

| Genes | Concordance, %<sup>†</sup> | Discordant Diplotypes |  |  |
| --- | --- | --- | --- | --- |
|  |  | Agena VeriDose® Core and *CYP2D6* CNV Panel | Nala PGx Core™ | Instances |
| *CYP2C9* | 100% (n = 221) | NA | NA | NA |
| *CYP2C19* | 96.4% (n = 223) | *1/*1 | *1/*3 | 1 |
|  |  | *1/*2 | *1/*1 | 1 |
|  |  | *1/*2 | *1/*3 | 1 |
|  |  | *1/*3 | *2/*2 | 1 |
|  |  | *2/*2 | *1/*1 | 2 |
|  |  | *2/*2 | *1/*2 | 2 |
| *CYP2D6* | 94.7% (n = 209) | *1/*1 | *2/*2 | 1 |
|  |  | *1/*10 | *1/*10, CN >=3 | 1 |
|  |  | *1/*41 | *39/*41 | 1 |
|  |  | *2/*2 | *1/*1 | 1 |
|  |  | *2/*10 | *1/*10 | 1 |
|  |  | *2/*36, CN >=3 | *2/*10, CN >=3 | 1 |
|  |  | *4/*36 | *4/*10 | 2 |
|  |  | *10/*36 | *2/*36 | 1 |
|  |  | *13 *1/*10 | *1/*10, CN >=3 | 1 |
|  |  | *13 *1/*41, CN = 1 | *1/*41 | 1 |

<sup>†</sup>The concordance presented in this table excludes samples that have mismatches in diplotype calls arising from technological differences between platforms. Please refer to Supplementary Table 7 for a detailed breakdown of all diplotypes detected.

Overall, a percentage agreement of 100% for *CYP2C9* (n=221), 96.4% for *CYPC219* (n=223) and 94.7% for *CYP2D6* (n=209) was observed between Nala PGx Core™ and the benchmark. Discordance was observed at n=1 for all diplotypes listed in Table 8 except for the following with more than one discordant calls: *CYP2C19* *2/*2 (n=4), and *CYP2D6* *4/*36 (n=2).

### Frequencies By Ethnicity

For samples that were concordant on Nala PGx Core™ and the benchmark platforms, we were able to observe the allele frequencies amongst the populations residing in Singapore and Indonesia (Table 9). From the combination of alleles present in individual’s chromosome, we were able to observe both the diplotype and corresponding phenotype frequencies amongst our study population (Table 10).

**Table 9.** Observed allele frequencies by ethnicity

| Gene | Allele or Variant | Allele Frequencies (Per This Study) |  |  |  |  |  | Allele Frequencies (PharmGKB) |  |  |
| --- | --- | --- | --- | --- | --- | --- | --- | --- | --- | --- |
|  |  | Indonesian | Chinese | Malay | Indian | Caucasian | Overall | East Asian | Central/South Asian | European |
| *CYP2C9*<br>(n = 206) | *2 | 0.000 | 0.000 | 0.000 | 0.060 | 0.172 | 0.032 | 0.002 | 0.114 | 0.127 |
|  | *3 | 0.000 | 0.040 | 0.039 | 0.100 | 0.069 | 0.044 | 0.038 | 0.110 | 0.076 |
| *CYP2C19*<br>(n = 201) | *2 | 0.297 | 0.274 | 0.289 | 0.229 | 0.138 | 0.256 | 0.284 | 0.270 | 0.147 |
|  | *3 | 0.041 | 0.055 | 0.066 | 0.000 | 0.017 | 0.042 | 0.072 | 0.016 | 0.002 |
|  | *17 | 0.054 | 0.007 | 0.039 | 0.229 | 0.138 | 0.067 | 0.021 | 0.171 | 0.216 |
| *CYP2D6*<br>(n = 195) | *2 | 0.088 | 0.118 | 0.167 | 0.140 | 0.196 | 0.136 | 0.121 | 0.295 | 0.277 |
|  | *4 | 0.000 | 0.035 | 0.069 | 0.020 | 0.161 | 0.051 | 0.005 | 0.091 | 0.185 |
|  | *5 | 0.029 | 0.028 | 0.028 | 0.080 | 0.000 | 0.031 | 0.049 | 0.046 | 0.030 |
|  | *6 | 0.000 | 0.000 | 0.000 | 0.000 | 0.018 | 0.003 | 0.000 | 0.000 | 0.011 |
|  | *9 | 0.000 | 0.000 | 0.000 | 0.000 | 0.018 | 0.003 | 0.002 | 0.003 | 0.028 |
|  | *10 | 0.338 | 0.299 | 0.222 | 0.160 | 0.143 | 0.251 | 0.436 | 0.087 | 0.016 |
|  | *14 | 0.000 | 0.014 | 0.000 | 0.000 | 0.000 | 0.005 | 0.003 | ND | 0.000 |
|  | *29 | 0.000 | 0.000 | 0.000 | 0.000 | 0.018 | 0.003 | 0.000 | 0.003 | 0.001 |
|  | *36 | 0.206 | 0.250 | 0.181 | 0.160 | 0.054 | 0.190 | 0.012 | 0.000 | 0.000 |
|  | *41 | 0.044 | 0.056 | 0.028 | 0.040 | 0.071 | 0.049 | 0.023 | 0.123 | 0.092 |
| *SLCO1B1*<br>(n = 203) | rs4149056<sup>‡</sup> | 0.125 | 0.074 | 0.026 | 0.040 | 0.167 | 0.084 | 0.125<sup>‡</sup> | 0.050<sup>‡</sup> | 0.159<sup>‡</sup> |

"ND" refers to instances in which no data is available for the given allele on PharmGKB.
<sup>‡</sup>rs4149056 refers to the reduced function variant of *SLCO1B1* that is present in *SLCO1B1*\*5, *SLCO1B1*\*15 and *SLCO1B1*\*17.
<sup>†</sup>Allele frequency values for rs4149056 have been obtained from gnomAD.

**Table 10.**
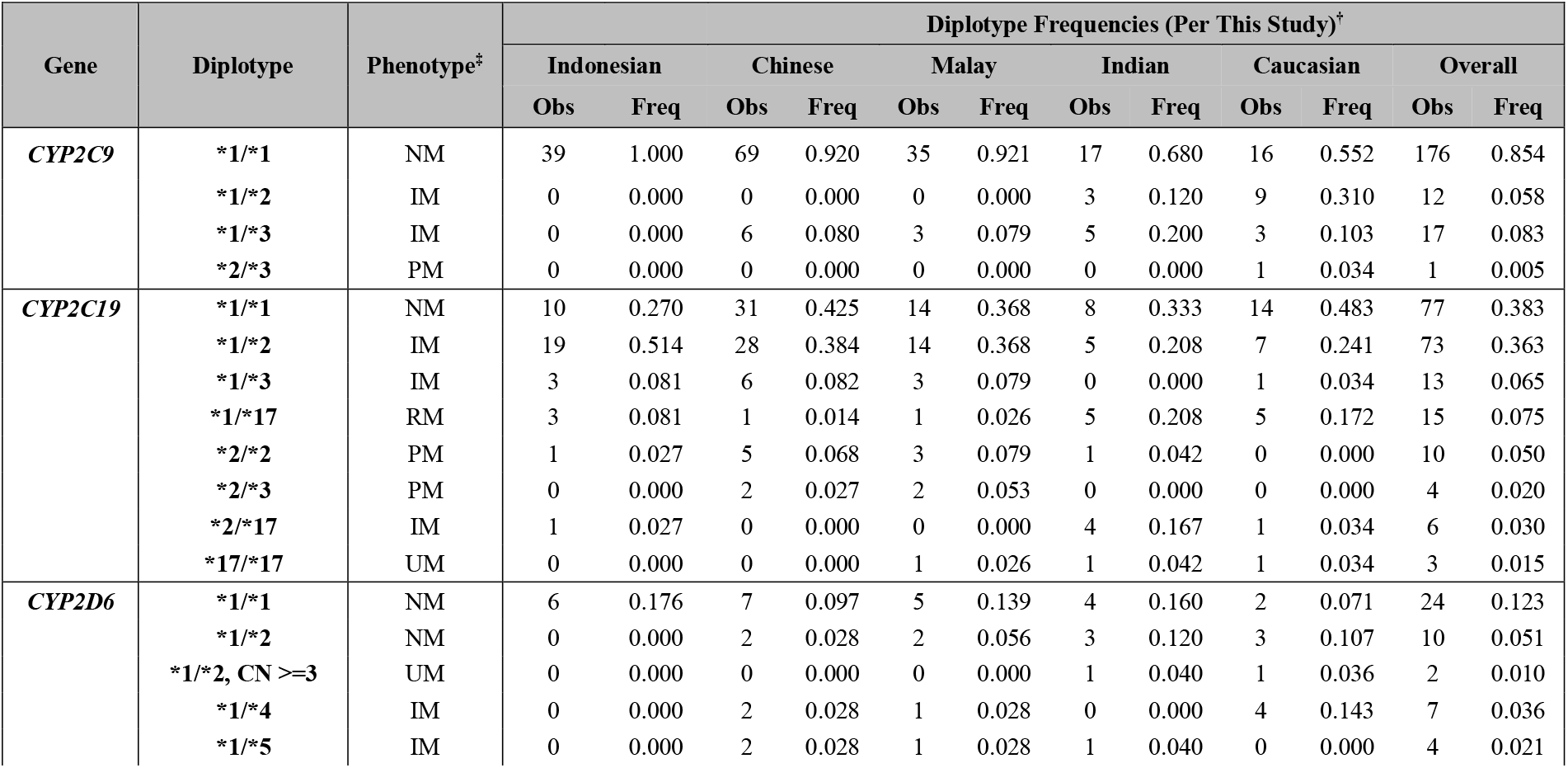
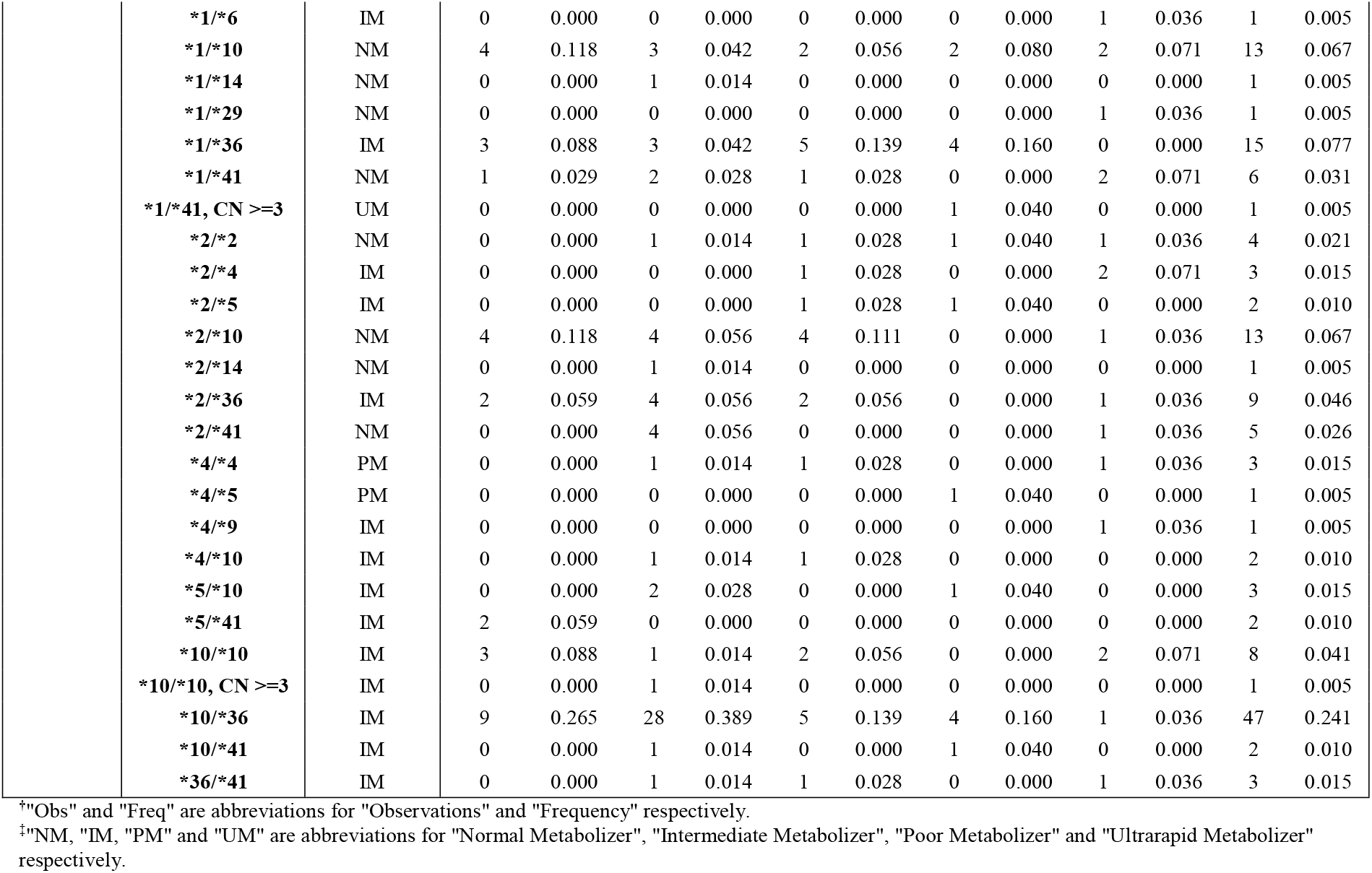
Observed diplotype frequencies by ethnicity

For *CYP2C9*, *3 allele was the most common amongst Chinese and Malay, and *2 allele amongst Caucasian, which is in line with PharmGKB’s reported distribution for the East Asian and European populations respectively. Our study also reported *3 allele as the more common variant in Indian population than *2, as opposed to PharmGKB’s frequency. These allele frequencies translated to *1/*3 as a common diplotype observed in Chinese, Malay and Indians, and *1/*2 in Caucasians.

For *CYP2C19*, we observed highest frequency of *CYP2C19*2* amongst Chinese, Malay and Indonesian which were categorized as East Asian populations. This resulted into high frequency of *1/*2 heterozygous depicted as a common diplotype amongst the population. The alleles *2 and *17 were observed as the common variants at equal proportions of 0.229 in Indians and 0.138 in Caucasians. *CYP2C19*3* was a common minor allele least observed amongst Indians and Caucasians, 0 and 0.017 respectively. As a result, *1/*2 and *1/*17 were common diplotypes observed in Indian and Caucasian populations, and *2/*17 only seen in Indians.

Common polymorphisms of *CYP2D6* in our population were seen in *10 and *36 alleles, at almost two-fold higher frequencies in Chinese, Malay and Indonesian than Indians. We noticed high frequencies of *36 in our East Asian population. Additionally, two functional copies of *36 allele were present in 1.4% of Chinese population who participated in our study (Figure 1). These alleles resulted in high frequency of *10/*36 as common diplotype amongst Chinese, Malay and Indonesian population. Our Indian population observed equal distribution of alleles *2, *10 and *36, although PharmGKB reported only *2 as the highest frequency in South Asians. The corresponding common diplotypes observed in Indians were of equal proportions in *1/*2, *1/*36 and *1/*10 ranging from 0.12 to 0.16. The alleles *2 and *4 were most common amongst Caucasians as reported by PharmGKB in Europeans, resulting in common diplotypes of *1/*2 and *1/*4 in the population.

**Figure 1.**
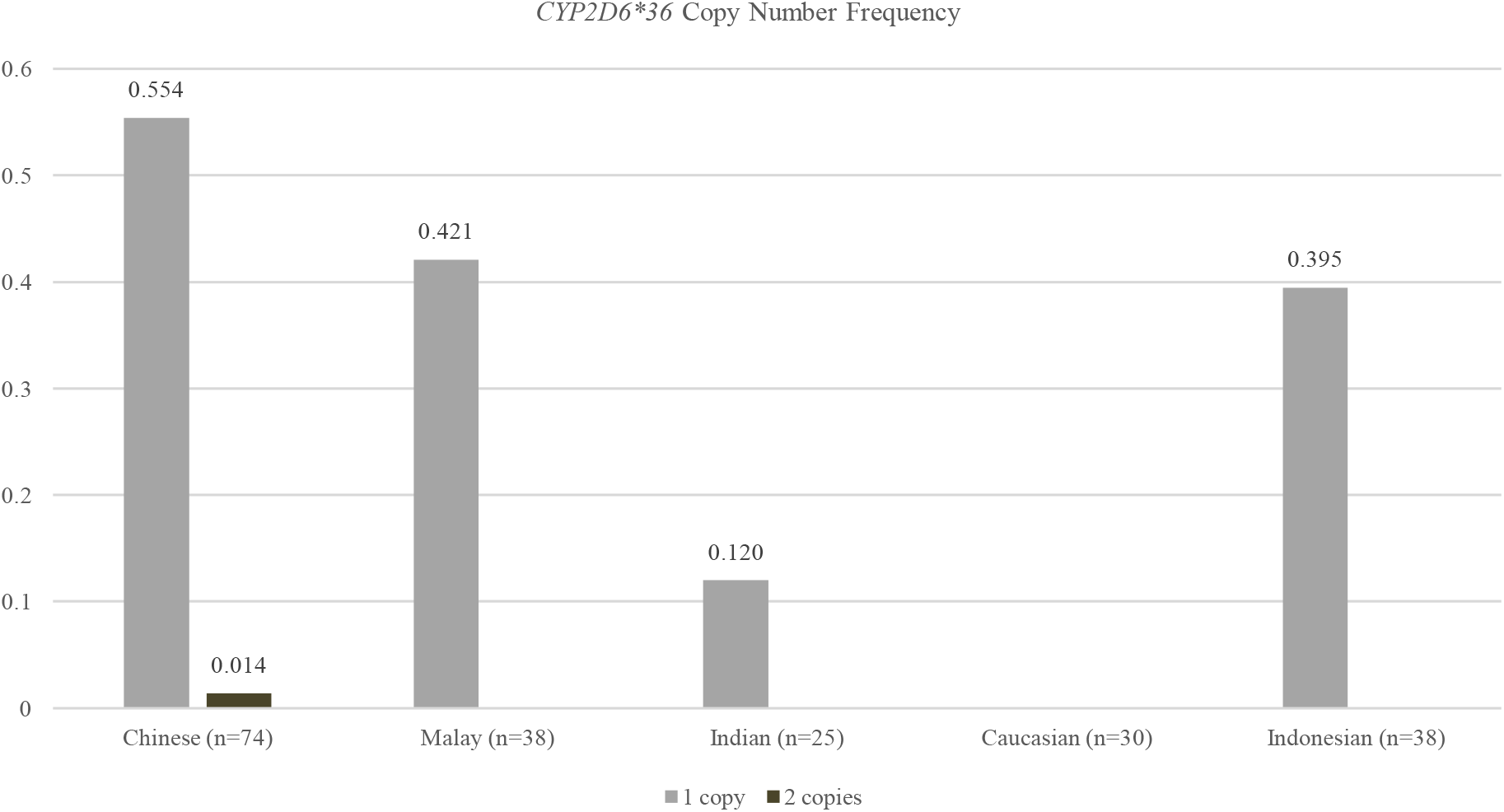
*CYP2D6*36* Frequency By Ethnicity. Distribution of individuals carrying one or two copies of the *CYP2D6*36* allele among the study cohort (n=205), grouped per ethnicity.

For *SLCO1B1*, the frequencies of rs4149056 across all ethnicities were consistent with values reported in gnomAD, with the variant being most common amongst Caucasians (0.167) and least amongst Indians (0.040). The frequency amongst East Asians (0.125, gnomAD), as denoted by the Chinese and Indonesian ethnic groups in this study, ranged between 0.074 and 0.125 respectively.

## Discussion

In this study, we evaluated the performance of Nala PGx Core™, a qPCR-based panel that evaluates 18 variants and 2 *CYP2D6* Copy Number markers across 4 pharmacogenes with established relevance across major ethnic groups in Singapore and Indonesia population. Nala PGx Core™ comes coupled with a reporting software that supports variant detection, diplotype assignment, diplotype-to-phenotype translation and the generation of reports containing clinical recommendations for each phenotype. The panel demonstrated high genotype-level call rates of >97% for *CYP2D6*, and 100% for *CYP2C9, CYP2C19* AND *SLCO1B1*. Similarly, high diplotype-level call rates were observed at >95% for *CYP2D6*, and 100% for *CYP2C9* and *CYP2C19*. A precision of 100% was observed under the same conditions (intra) and across different conditions (inter). In comparison to other established platforms serving as benchmarks during the study, Nala PGx Core™ had ≥96.9% concordance rate for all variant level assays, which consequently resulted in ≥94.7% concordance at a diplotype level across *CYP2C9, CYP2C19* and *CYP2D6*.

Failures to produce a variant genotype call could be attributed to several reasons. Firstly, failures could potentially stem from the quality of gDNA, despite the DNA quality checks (QC) performed prior to accepting a sample for testing. Poor DNA quality could arise from multiple factors along the sample handling chain. Such factors include the contamination of the buccal fluid by interfering particles during sample collection, inconsistent conditions during sample transport and human error during sample purification. These may lead to the degradation of genomic DNA, poor homogenization of the sample in collection and/or extraction buffers, and the carryover of contaminants, thereby compromising sample integrity^32^. Further QC that involves specific quantification of double-stranded non-fragmented DNA and traces of other interfering materials like RNA, carryover carbohydrate, residual phenol, guanidine or other reagents could enhance the call rate^33^. Regardless, the overall higher variant call rates on Nala PGx Core™ panel demonstrate high tolerance of interfering substances, therefore alluding to the high robustness of the assay. Often, failures at variant genotyping subsequently contribute to failures at determining diplotype, since an incomplete variant panel cannot translate into a diplotype. Failures at diplotype calling could also arise from a combination of variants that do not map onto a distinct diplotype, per the reference database, potentially indicating a novel combination.

We further sought to evaluate the allele frequency distribution in the study cohort across the 5 major ethnic groups observed (Indonesian, Chinese, Malay, Indian and Caucasian). The data presented was limited strictly to the geographical boundaries of Singapore and Indonesia, which could account for the difference in allele frequencies observed in comparison to PharmGKB, which is representative of a more expansive and global cohort^34^. Whilst dissimilar to database figures, our study demonstrated the distributions for the following to be concordant with previous studies^7,35,36^, suggesting a niche in the PGx landscape of Singapore and Indonesia –

1. *CYP2C9*3* allele as the more common variant within Indians than *CYP2C9*2*^7^
2. A *SLCO1B1* rs4149056 frequency of 12.5% amongst Indonesians^35^
3. High frequencies of the *CYP2D6*10* amongst Indonesians^35^
4. High frequencies of the *CYP2D6*36* allele as seen in the Indonesian, Chinese and Malay ethnicities^36^
5. Two functional copies of *CYP2D6* within our Chinese population^36^

Due to the lack of *CYP2D6* copy number references^35^, it is our understanding that the frequency of *CYP2D6*36* in Indonesia may not be well-represented. Our study revealed that the prevalence of *CYP2D6*36* to be approximately seventeen times higher amongst Indonesians as compared to the corresponding East Asian allele frequency on PharmGKB^37^. Furthermore, our study provides insight on the frequency of the *CYP2D6* *10/*36 diplotype in the archipelago, which may help inform the adoption of population-specific PGx workflows in the region. Taken together, the data presents a case for extending tailored PGx testing across the 4 pharmacogenes studied, *CYP2C9, CYP2C19, CYP2D6* and *SLCO1B1*, in South East Asia.

### Limitations

The single largest reason for the diplotype-level discordance observed between Nala PGx Core™ and the Agena VeriDose^®^ Core and *CYP2D6* CNV Panel appears to be a difference in the technological offerings and hence, allele coverage of each panel. From Supplementary Table 7, out of the 28 instances of discordance observed, 9 instances were attributed to this cause (32.1%). This included two alleles each in *CYP2C9, CYP2C19* and *CYP2D6*.

The variants *CYP2C9*12, CYP2C9*13, CYP2C19*4A, CYP2C19*6* and *CYP2D6*13* were covered by Agena VeriDose^®^ Core but not by Nala PGx Core™. Due to lack of coverage for these alleles, the genotypes identified by Nala PGx Core™ were inconsistent with the calls produced by Agena VeriDose^®^ Core. It is important to note that Nala PGx Core™ was curated specifically to include alleles that were relevant to Asian and European populations, with a Minor Allele Frequency (MAF) >1% based on PharmGKB database for East Asians, South Asians and Europeans^37^. Similarly, *CYP2D6*35*, an allele present in 5.5% of Europeans and 1.2% of South Asians, was identified by Nala PGx Core™ but not by Agena VeriDose^®^ Core. The technological differences and subsequent inconsistencies in reporting suggest the need for further review from a product development standpoint.

For the highly polymorphic *CYP2D6* gene, Nala PGx Core™ was designed to only cover the variant alleles listed in Table 1, that could lead to overestimation of *1 allele in our population. This means star alleles that may be present in the population were identified as normal genotype, which could result in overestimation of *1 allele in our population as compared to the PharmGKB’s reported frequency (Supplementary Table 4).

The study also relied on self-reported ethnicity, which may have contributed to racial bias in summarizing the observed allele and diplotype frequencies per ethnic group. Future studies may consider screening the ethnicities of participants as well as their next-of-kin, up to two generations prior^10^, to improve classification accuracy.

Finally, the study cohort consisted of 246 individuals, with each ethnic group consisting of approximately 24 to 75 individuals for the frequency analysis. Larger scale studies may be conducted to ensure that the trends observed as part of this study may be generalized across the populations examined.

### Future Work

Further investigation is required for the remaining samples that had discrepant genotypes across platforms. Nala PGx Core™ was designed as a qPCR platform and genotyping was accompanied by Nala CDS™ that utilized quantitative thresholds. There could be cases of samples displaying borderline or ambiguous qPCR amplification curves and Ct-values, which could be misinterpreted by these thresholds. The benchmark platforms, Agena VeriDose^®^ Core and *CYP2D6* CNV Panel and TaqMan^®^ DME Genotyping Assays, were also analyzed by their accompanying software packages. Such ambiguous cases on either platform could lead to instances of discordance and sequencing of the region of interest may be required to ascertain the truth. For gene variants caused by SNPs and indels, the qPCR amplified product can be submitted for genome sequencing^13^. In the investigation of copy number variants, a two-step approach can be adopted^38^. Firstly, CNV calls observed in this study may be checked against an alternative benchmark such as digital droplet PCR (ddPCR)^39^. Secondly, where discordance in CNV calling persists, methods such as XL-PCR can be adopted to identify the presence of *CYP2D6/CYP2D7* hybrid states, with submitting samples for whole-genome sequencing^38^ being an option. Investigation of copy number variants may require amplification of region spanning across the gene of interest.

## Data Availability

The data that supports the findings of this study are available as supplementary materials. Any additional data is available from the corresponding author upon reasonable request.

## Acknowledgements

We would like to thank Mr. Xie Zhicheng and Mr. Heng Khai Koon at the Genome Institute of Singapore for their assistance with performing the Agena VeriDose^®^ Core and *CYP2D6* CNV Panel studies. We also thank PT Nalagenetik Riset Indonesia for their assistance in study recruitment and experimentation performed in Indonesia, and Genome Institute of Singapore for providing facility to perform all experimentations done in Singapore.

## Supplementary Tables And Figure Legends

**Supplementary Table 1**. Observed Genotype-Level Call Rates Per Variant Per Gene Per Platform, With Counts Of Successful And No Calls

**Supplementary Table 2**. Observed Diplotype-Level Call Rates Per Gene Per Platform, With Counts Of Successful And No Calls

**Supplementary Table 3**. Precision Analysis Of Nala PGx Core™

**Supplementary Table 4**. Observed allele frequencies by ethnicity

**Supplementary Table 5**. *CYP2D6* Intron 2 And Exon 9 Observations And Frequencies By Ethnicity

**Supplementary Table 6**. *CYP2D6*36* Observations And Frequencies By Ethnicity

**Supplementary Table 7**. Diplotype Calls Observed Across All Samples Tested On Agena Veridose^®^ Core And *CYP2D6* CNV Panel And Nala PGx Core™

**Supplementary Table 8**. Raw And Reported Diplotype Concordance Values For Samples Tested On Agena Veridose^®^ Core And CYP2D6 CNV Panel And Nala PGx Core™

**Supplementary Tables 9.1 to 9.18**. Raw genotype calls for rs1799853, rs1057910, rs4244285, rs4986893, rs12248560, rs1065852, rs5030655, rs3892097, rs35742686, rs16947, rs28371725, rs1135840, rs769258, rs5030865, rs5030656, rs59421388 and rs267608319, on Nala PGx Core™ and the corresponding benchmark methods, Agena VeriDose^®^ Core and *CYP2D6* CNV Panel or TaqMan^®^ DME Genotyping Assays

**Supplementary Table 9.19**. Raw variant calls for *CYP2D6* Intron 2 and Exon 9 on Agena VeriDose^®^ Core and *CYP2D6* CNV Panel and Nala PGx Core™

**Supplementary Figure 1**. Observed *CYP2D6* Functional Copy Number Frequencies By Ethnicity. Distribution of individuals carrying one, two or three or more copies of the *CYP2D6* functional gene among the study cohort (n=205), grouped per ethnicity.

## References

1. Berm EJJ, Looff M de, Wilffert B, et al. Economic Evaluations of Pharmacogenetic and Pharmacogenomic Screening Tests: A Systematic Review. Second Update of the Literature. Bruns H, ed. PLoS One. 2016;11(1):e0146262. doi:10.1371/journal.pone.0146262

2. Chan SL, Ang X, Sani LL, et al. Prevalence and characteristics of adverse drug reactions at admission to hospital: a prospective observational study. Br J Clin Pharmacol. 2016;82(6):1636–1646. doi:10.1111/bcp.13081

3. Sultana J, Cutroneo P, Trifirò G. Clinical and economic burden of adverse drug reactions. J Pharmacol Pharmacother. 2013;4(SUPPL.1). doi:10.4103/0976-500X.120957

4. Zhu Y, Lopes GS, Bielinski SJ, et al. Impact of Pharmacogenomic Information on Values of Care and Quality of Life Associated with Codeine and Tramadol-Related Adverse Drug Events. Mayo Clin Proc Innov Qual Outcomes. 2021;5(1):35–45. doi:10.1016/j.mayocpiqo.2020.08.009

5. Furge LL, Guengerich FP. Cytochrome P450 enzymes in drug metabolism and chemical toxicology: An introduction. Biochem Mol Biol Educ. 2006;34(2):66–74. doi:10.1002/bmb.2006.49403402066

6. Malsagova KA, Butkova T V, Kopylov AT, et al. Pharmacogenetic Testing: A Tool for Personalized Drug Therapy Optimization. Pharmaceutics. 2020;12(12):1–23. doi:10.3390/pharmaceutics12121240

7. Ling Goh L, Wei Lim C, Cheng Sim W, Xian Toh L, Pang Leong K. Analysis of Genetic Variation in CYP450 Genes for Clinical Implementation. 2017. doi:10.1371/journal.pone.0169233

8. Kisor DF, Hoefer C, Decker BS. Pharmacogenomics and precision medicine. In: Clinical Pharmacy Education, Practice and Research: Clinical Pharmacy, Drug Information, Pharmacovigilance, Pharmacoeconomics and Clinical Research. Elsevier; 2018:437–451. doi:10.1016/B978-0-12-814276-9.00031-3

9. Ooi BNS, Raechell Ying AF, et al. Robust Performance of Potentially Functional SNPs in Machine Learning Models for the Prediction of Atorvastatin-Induced Myalgia. Front Pharmacol. 2021;12:605764. doi:10.3389/fphar.2021.605764

10. Brunham LR, Chan SL, Li R, et al. Pharmacogenomic diversity in Singaporean populations and Europeans. Pharmacogenomics J. 2014;14(6):555–563. doi:10.1038/tpj.2014.22

11. Bachtiar M, Ooi BNS, Wang J, et al. Towards precision medicine: interrogating the human genome to identify drug pathways associated with potentially functional, population-differentiated polymorphisms. Pharmacogenomics J. 2019;19(6):516–527. doi:10.1038/s41397-019-0096-y

12. Perkel J. SNP genotyping: Six technologies that keyed a revolution. Nat Methods. 2008;5(5):447–453. doi:10.1038/nmeth0508-447

13. Yang W, Wu G, Broeckel U, et al. Comparison of genome sequencing and clinical genotyping for pharmacogenes. Clin Pharmacol Ther. 2016;100(4):380–388. doi:10.1002/cpt.411

14. S.A. Deepak, K.R. Kottapalli, R. Rakwal, et al. Real-Time PCR: Revolutionizing Detection and Expression Analysis of Genes. Curr Genomics. 2007;8(4):234–251. doi:10.2174/138920207781386960

15. Pratt VM, Zehnbauer B, Wilson JA, et al. Characterization of 107 genomic DNA reference materials for CYP2D6, CYP2C19, CYP2C9, VKORC1, and UGT1A1: A GeT-RM and association for molecular pathology collaborative project. J Mol Diagnostics. 2010;12(6):835–846. doi:10.2353/jmoldx.2010.100090

16. Pratt VM, Everts RE, Aggarwal P, et al. Characterization of 137 Genomic DNA Reference Materials for 28 Pharmacogenetic Genes: A GeT-RM Collaborative Project. J Mol Diagnostics. 2016;18(1):109–123. doi:10.1016/j.jmoldx.2015.08.005

17. Syvänen AC. Accessing genetic variation: Genotyping single nucleotide polymorphisms. Nat Rev Genet. 2001;2(12):930–942. doi:10.1038/35103535

18. Suo W, Shi X, Xu S, Li X, Lin Y. Towards low cost, multiplex clinical genotyping: 4-fluorescent Kompetitive Allele-Specific PCR and its application on pharmacogenetics. Kalendar R, ed. PLoS One. 2020;15(3):e0230445. doi:10.1371/journal.pone.0230445

19. Shen GQ, Abdullah KG, Wang QK. The TaqMan method for SNP genotyping. Methods Mol Biol. 2009;578:293–306. doi:10.1007/978-1-60327-411-1_19

20. Davis BH, DeFrank G, Limdi NA, Harada S. Validation of the Spartan RX CYP2C19 Genotyping Assay Utilizing Blood Samples. Clin Transl Sci. 2020;13(2):260–264. doi:10.1111/cts.12714

21. Zhou Y, Armstead AR, Coshatt GM, Limdi NA, Harada S. Comparison of two point-of-care cyp2c19 genotyping assays for genotype-guided antiplatelet therapy. Ann Clin Lab Sci. 2017;47(6):738–743.

22. Bergmeijer TO, Vos GJ, Claassens DM, et al. Feasibility and implementation of CYP2C19 genotyping in patients using antiplatelet therapy. Pharmacogenomics. 2018;19(7):621–628. doi:10.2217/pgs-2018-0013

23. Chae H, Kim M, Koh Y-S, et al. Feasibility of a Microarray-Based Point-of-Care CYP2C19 Genotyping Test for Predicting Clopidogrel On-Treatment Platelet Reactivity. Biomed Res Int. 2013;2013:1–5. doi:10.1155/2013/154073

24. Lee CC, McMillin GA, Babic N, Melis R, Yeo K-TJ. Evaluation of a CYP2C19 genotype panel on the GenMark eSensor® platform and the comparison to the Autogenomics Infiniti™ and Luminex CYP2C19 panels. Clin Chim Acta. 2011;412(11-12):1133–1137. doi:10.1016/j.cca.2011.03.001

25. Vanwong N, Ngamsamut N, Hongkaew Y, et al. Detection of CYP2D6 polymorphism using Luminex xTAG technology in autism spectrum disorder: CYP2D6 activity score and its association with risperidone levels. Drug Metab Pharmacokinet. 2016;31(2):156–162. doi:10.1016/j.dmpk.2016.01.005

26. Lois A, Everts RE, Nakorchevsky A, et al. The VeriDose Core Panel: Strong Performance When Analyzing Challenging Pharmacogenetic Samples. https://agenabio.com/wp-content/uploads/2020/06/Agena-Conference-Poster-Veridose-Core-V3-G020.pdf. Accessed April 2, 2021.

27. Gaedigk A, Turner A, Everts RE, et al. Characterization of Reference Materials for Genetic Testing of CYP2D6 Alleles. J Mol Diagnostics. 2019;21(6):1034–1052. doi:10.1016/j.jmoldx.2019.06.007

28. Technologies L. TaqMan ® SNP Genotyping Assays TaqMan ® Predesigned SNP Genotyping Assays, TaqMan ® Custom SNP Genotyping Assays, and TaqMan ® Drug Metabolism Enzyme Genotyping Assays.; 2014.

29. Population Genetics and the Hardy-Weinberg Principle. https://www.cs.cmu.edu/~genetics/units/instructions/instructions-PGE.pdf. Accessed May 2, 2021.

30. Allele frequency. https://www.nature.com/scitable/definition/allele-frequency-298/. Accessed May 2, 2021.

31. Calculating Inter- and Intra-Assay Coefficients of Variability. https://salimetrics.com/calculating-inter-and-intra-assay-coefficients-of-variability/. Accessed May 1, 2021.

32. Genomic DNA Preparation Troubleshooting. https://www.sigmaaldrich.com/ID/en/technical-documents/technical-article/genomics/dna-and-rna-purification/problems-during-genomic-dna-preparation. Accessed June 30, 2021.

33. Matlock B. Assessment of Nucleic Acid Purity. Tech Bull NanoDrop Spectrophotometers. 2015:1–2.

34. Huddart R, Fohner AE, Whirl□Carrillo M, et al. Standardized Biogeographic Grouping System for Annotating Populations in Pharmacogenetic Research. Clin Pharmacol Ther. 2019;105(5):1256–1262. doi:10.1002/cpt.1322

35. Runcharoen C, Fukunaga K, Sensorn I, et al. Prevalence of pharmacogenomic variants in 100 pharmacogenes among Southeast Asian populations under the collaboration of the Southeast Asian Pharmacogenomics Research Network (SEAPharm). Hum Genome Var. 2021;8(1):7. doi:10.1038/s41439-021-00135-z

36. Chan W, Li MS, Sundaram SK, Tomlinson B, Cheung PY, Tzang CH. CYP2D6 allele frequencies, copy number variants, and tandems in the population of Hong Kong. J Clin Lab Anal. 2019;33(1). doi:10.1002/jcla.22634

37. Whirl-Carrillo M, McDonagh EM, Hebert JM, et al. Pharmacogenomics Knowledge for Personalized Medicine. Clin Pharmacol Ther. 2012;92(4):414–417. doi:10.1038/clpt.2012.96

38. Pös O, Radvanszky J, Styk J, et al. Copy Number Variation: Methods and Clinical Applications. Appl Sci. 2021;11(2):819. doi:10.3390/app11020819

39. Gross AM, Ajay SS, Rajan V, et al. Copy-number variants in clinical genome sequencing: deployment and interpretation for rare and undiagnosed disease. Genet Med. 2019;21(5):1121–1130. doi:10.1038/s41436-018-0295-y

